# The relationships between experimental task and questionnaire measures of reward/punishment sensitivity in attention deficit hyperactivity disorder (ADHD): protocol for a scoping review

**DOI:** 10.1101/2023.07.21.23292991

**Authors:** Mana Oguchi, Emi Furukawa, Naano Nagahama, Kokila Dilhani Perera, Gail Tripp

## Abstract

**Introduction:** One of the purported underlying causal mechanisms of attention deficit hyperactivity disorder (ADHD) is altered motivational processes. Questionnaires have been used to identify the characteristics of reward and punishment sensitivity in individuals with ADHD. However, these questionnaires were initially developed to measure individual traits related to anxiety (inhibitory) and impulsivity (approach) tendencies or differences in pleasure-seeking. These reward and punishment sensitivity questionnaires are useful but might not capture all relevant aspects of altered motivational processes in ADHD. The proposed scoping review aims to: 1) examine which aspects of hypothesized altered reward and punishment sensitivity correspond to constructs measured by existing questionnaires, 2) characterize the relationships between ADHD symptomatology and reward and punishment sensitivity as measured by existing questionnaires, and 3) evaluate the consistency between the altered reward and punishment sensitivity as measured by existing questionnaires and experimental task performance.

**Methods and analysis:** This scoping review will adhere to the Preferred Reporting Items for Systematic Reviews and Meta-Analyses (PRISMA) Extension for Scoping Reviews and the Joanna Briggs Methodology for Scoping Reviews. Published English language literature will be searched in three electronic databases, with no restriction on the year of publication. Two researchers will independently screen all identified titles/abstracts and review the method sections of the identified papers to confirm their eligibility before proceeding to full-text review and data extraction. Methods, results, and conclusions will be tabulated by research questions. A narrative review, and summary conclusions will be presented. The evidence will be summarized as descriptive data in the Excel table.

**Ethics and dissemination:** This study reviews existing publications with ethical approval in place. Therefore, ethical approval is not required. Review results will be disseminated through academic conferences and peer-reviewed manuscripts. Scoping review results will also inform future research to measure and identify altered motivational processes in ADHD.

**Strengths and limitations of this study:**

- This scoping review is the first study to identify which aspects of ADHD reinforcement sensitivity have been measured by existing reward and punishment scales and comprehensively review studies reporting relationships between experimental task results and reward and punishment scales in ADHD. (strengths)
- This study will be conducted according to PRISMA guidelines for systematic reviews. (strength)
- Results will be summarized separately for children/adolescents and adults. (strength)
- This review includes only published peer-reviewed English language studies. (limitation)

## INTRODUCTION

### Review questions

ADHD is a common neurodevelopmental disorder characterized by three cardinal symptoms: inattention, hyperactivity, and impulsivity [1]. The prevalence of ADHD is approximately 7.6 % in children and 2.6% in adults [2, 3], with some symptom fluctuation across the lifespan [3–5]. Altered motivational processes have been proposed to account for symptoms of ADHD [6–9]. Behavioral studies have identified altered sensitivity to both reward and punishment in individuals with ADHD [10]. Compared to typically developing peers, children with ADHD have been shown to prefer immediate over delayed reward [11–13], to show poorer adaptation to changing reinforcement contingencies [14, 15], and demonstrate faster extinction after learning under partial (discontinuous) reinforcement [16]. There have been fewer studies of sensitivity to punishment in those with ADHD, and the results are mixed. Some studies have shown similar sensitivity to punishment between those with and without ADHD [17, 18], while others report increased sensitivity to punishment amongst those with ADHD [15, 19].

While most of the evidence on altered motivational processing in ADHD comes from experimental studies, questionnaires assessing sensitivity to reward and punishment have also been used. The only questionnaire developed specifically to assess for altered reward sensitivity is the Quick Delay Questionnaire, designed for use with adults, which assesses feelings/attitudes toward waiting and delayed rewards [20]. Individuals with ADHD report higher levels of delay aversion and delay discounting [21], compared to their typically developing peers, which is consistent with the available experimental findings [11, 22].

Other reward and punishment sensitivity questionnaires have been developed for other pathological conditions or are based on reinforcement learning theories. Studies using these questionnaires report inconsistent results in terms of reward and punishment sensitivity in ADHD [23–25]. It is unclear whether they are measuring the same motivational constructs as those evaluated in experimental studies. The most commonly used questionnaires on reward and punishment sensitivity [26] were developed based on Gray’s Reinforcement Sensitivity Theory (RST) [27]. This theory conceptualizes reward sensitivity as a biologically based behavioral activation/approach system (BAS), i.e., a temperamental trait to seek rewarding stimuli. Punishment sensitivity is thought to relate to the behavioral inhibition system (BIS), i.e., an anxiolytic trait to avoid potentially aversive stimuli [28]. An unbalanced BIS/BAS has been linked to increased risks of psychopathology [29], including ADHD [30, 31]. Quay [32] assumed an underactive BIS leads to an inhibition deficit, poor attention and stimulus seeking in ADHD, while other researchers demonstrated that a high or a dysregulated BAS underlies elevated levels of hyperactive and impulsive behaviors [33–35].

Other questionnaires measure reward anticipation (‘wanting’) and consumption (‘liking’) as expressed in behavior [36] and urges/pleasure seeking [37]. Using a range of questionnaires, excessive, or reduced, reward seeking, anticipation, and consumption have been linked with a range of pathological conditions, including addiction [38, 39], eating disorders [40], and anhedonia [41, 42]. Links between the constructs measured by these questionnaires and ADHD are unclear. Using the UPPS Impulsive Behavior Scale [43, 44] some studies have reported greater urgency to obtain rewards in those with ADHD [45, 46]. Other measures have been developed to examine anhedonia. Many of these measures examine behavior or mood symptoms associated with specific disorders (e.g., depression) or have items that name specific reward stimuli (e.g., social, food) [41, 47–49]. In a study using the Tripartite Pleasure Inventory, Meinzer and colleagues [50] suggested that a reduced capacity to attend to pleasurable stimuli/experiences led to a disorganized pursuit of rewards in those with ADHD.

A smaller number of questionnaires are available to assess sensitivity to punishment in addition to those developed based on, or elaborated from, the RST. Some of these measures attempted to better differentiate responsiveness to punishment and motivation to avoid punishment [51], or removed mention of specific aversive stimuli from questionnaire items (e.g., the Reward and Punishment Responsivity and Motivation Questionnaire) [51]. Using a measure of avoidance of negative outcomes (e.g., Acceptance and Action Questionnaire-□) Bond and colleagues [52] report that individuals with ADHD show increased avoidance of negative thoughts, feelings, and other internal experiences [53]. In children, symptoms of disruptive behavior disorders may imply reduce responsiveness to punishment, i.e., repetitive and persistent patterns of inappropriate behavior despite negative consequences. One questionnaire (Multidimensional Assessment Profile of Disruptive Behavior) [54] explicitly measures insensitivity to punishment. In this questionnaire, temper loss, irritability, and frustration are also conceptualized as overreactions to aversive stimuli/results or non-reward [55]. However, to our knowledge, the association between this questionnaire and ADHD symptoms has not been explored.

Despite the importance of motivational processes in identifying and describing the characteristics of ADHD, it is unclear whether experimental and questionnaire-based studies evaluate the same aspects of reward and punishment sensitivity or provide consensus. Therefore, this scoping review aims to answer the following questions:

1. Which aspects of hypothesized altered reward and punishment sensitivity in ADHD correspond to the constructs measured by existing questionnaires?
2. What are the relationships between ADHD symptomatology and reward and punishment sensitivity as measured by existing questionnaires?
3. What is the degree of consistency between the experimental and questionnaire findings on reward and punishment sensitivity in ADHD?

By addressing the above questions, this study will identify the overlap and differences in the measurement of reinforcement sensitivity by experimental tasks and questionnaires?

## METHODS AND ANALYSIS

This review protocol will follow the Joanna Briggs Institute (JBI) Methodology for Scoping Reviews [56, 57]. Further, this scoping review will be formatted along the PRISMA Extension for Scoping Reviews guidelines (PRISMA-ScR; supplement 1) [58]. The review will run from 17 July (estimated completion date of the search strategy), 2023, through 15 August 2023 (estimated completion date of the review extraction of data).

### Inclusion criteria

#### Participants

This scoping review will include studies with participants of any age who have a clinical diagnosis of ADHD or elevated ADHD symptoms as reported by parents and teachers in the case of children or adult self-report. Studies that evaluate other neuro-developmental or psychiatric disorders, and do not include ADHD-only groups, will be excluded. Where such studies include ADHD groups, only the results for ADHD will be reported. Studies focused on non-human participants will not be included.

#### Concept

This scoping review will focus on the motivational processes in ADHD and will examine questionnaires that measure reward and punishment sensitivity in ADHD and where applicable their relationship to the results of experimental studies. Specifically, this review will include two types of studies: (1) studies that describe ADHD symptoms/diagnostic status and measure and report reward and punishment sensitivity using questionnaires, and (2) studies that measure and report reward and punishment sensitivity by both questionnaires and experimental tasks. These studies will be organized separately for children/adolescents and adults. The following papers will not be included: case reports, reviews or systematic literature reviews, qualitative studies, opinion pieces, editorials, comments, news, letters to the editor that do not include empirical research, and non-human studies. However, in the Introduction and Discussion sections, the above literature may be reviewed and discussed.

#### Context

This study’s context will be open and will include all published studies (meeting the criteria for the above concept and participants) using questionnaire scales and experimental tasks evaluating reward and punishment sensitivity in ADHD. Furthermore, ADHD in this study includes the presence of an ADHD diagnosis as well as elevated symptoms of ADHD as defined by the reviewed manuscript authors (may include above- and sub-threshold levels of ADHD). Both clinical and community samples, and all geographic regions/settings, races, and genders will be included.

#### Types of evidence

Any study design that meets inclusion criteria, including self-report data, data obtained from parents, teachers, or researchers, and experimental tasks, will be included in the scope review. There are no restrictions on the year of publication. However, the search will be limited to full-text articles of primary research published in English in peer-reviewed journals. Thus, systematic reviews, meta-analyses, case reports, commentaries, posters, opinion pieces, editorials, comments, newsletters, letters to the editor with no empirical research, non-human studies, and gray literature will be excluded due to resource constraints and to be consistent with the purpose of this study.

#### Search strategy

This search will be conducted across three databases using the search engine Google: PubMed (MEDLINE), Web of Science, APA PsycINFO (Ovid). The following search terms and synonyms will be used: population (e.g., attention deficit disorder with hyperactivity [MeSH Terms]), measurements (e.g., surveys and questionnaires [MeSH Terms]), and the concepts of measurement (e.g., reinforcement, psychology [MeSH Terms], reward sensitivity [All Fields], punishment sensitivity [All Fields]). All phrases considered were included in the search string. Using several databases, preliminary searches were carried out to determine keywords, descriptors, and Medical Subject Headings (MeSH). The completed search strategy was built based on those searches, and the determined items were integrated with the Boolean operators AND, OR, and NOT. The draft version of the search strategy for the PubMed, Web of Science, and PsycInfo databases is included in supplementary appendix 2. This search was conducted on 17 July 2023 through those databases and 4,729 papers without duplicates were identified. We will find related measurement reports via different databases or sources (e.g., Google Scholar) and list all articles citing those papers. Searches from citations in papers that met the criteria will be included. The final scoping review will include the detailed search strategies for all sources.

#### Study selection

To validate the inclusion/exclusion criteria, preliminary searches were conducted on several databases. Based on the preliminary search, this review will be implemented using an integrated research strategy with Boolean operators AND and OR. The final search will take place between July to August, and hand searching will continue throughout in abstract review period. All studies identified by the database and hand search will be grouped and duplicates removed using Covidence software [59]. The software is a web platform tool with functions for systematic reviews, including importing and screening literature and assessing the risk of bias. Using this software, two independent researchers (MO and NN) will screen to verify the presence or absence of study eligibility criteria, participants, concept, and context. The researchers will first screen all identified titles/abstracts (ADHD or related terms and the use of a reward and punishment sensitivity questionnaire or experimental task must be mentioned) and the same researchers will then review the method sections of the papers to examine if studies meet the full inclusion criteria. All studies excluded during the screening phase and the reasons for exclusion will be reported. Any conflicts in both phases of screening (abstract and full-text screening) will be resolved through discussion between the two researchers and consultation with the research team (protocol authors).

#### Data extraction

The data included in the final paper of this scoping review will be extracted by two independent researchers (MO, NN) using a data extraction form developed based on the JBI template [60]. The template for data extraction may be further refined and updated throughout the review phase. Individual research methods, participants, measurements, results, and conclusions will be summarized in a table using an Excel file. If necessary, the authors of the published papers will be contacted to request missing or additional data. The study description and research questions will be retained as information of interest for this review (Supplementary appendix 3).

#### Presentation of results

All information regarding the selection of papers is presented in a flow figure according to PRISMA-ScR (Supplementary appendix 1). The results of all studies that meet the criteria will be summarized in tables with descriptive data and in narrative form. In addition, the tables of results will include correlation coefficients between experimental tasks and questionnaires where these are available. Evidence will be summarized as the number of papers and categorized by article type and study design type to highlight areas where additional research may be needed to fill current evidence gaps. This scoping review will identify gaps in what has been measured in terms of reward and punishment sensitivity in ADHD using existing questionnaires. The results will be reported separately for childhood/adolescents (under the age of 18) and adults (over 18) [61].

Implications for future research will be discussed based on the relationship between the findings from questionnaire research and hypothesized altered motivational processing as well as experimental research results. There will also be a discussion of the need and feasibility of future research on questionnaires in the context of ADHD motivational processing and a systematic review/meta-analysis of response differences in questionnaires by ADHD symptom status/severity.

#### Participants and public involvement

The protocol and scoping review will not include members of the general public or patients.

## Supporting information

PRISMA-ScR

## Data Availability

All data produced in the present study are available upon reasonable request to the authors.

## ETHICS AND DISSEMINATION

Since this proposed study is a scoping review, there are no required ethical or safety considerations. The results of the scoping review will be disseminated through publication in peer-reviewed journals and conference presentations.

## Acknowledgments

Nothing.

## Contributors

MO, EF, and GT contributed to the conception and design of this protocol. MO and NN led the search strategy and pilot data mapping, supported by EF, KD, and GT. MO drafted the manuscript and EF and GT contributed to the revisions. KD assessed this protocol along with the PRISMA-ScR checklist, which was reviewed by all authors. All authors (MO, EF, NN, KD, GT) approved the final version of the manuscript for accuracy, completeness, and publication.

## Patient consent for publication

Not required.

## Provenance and peer review

Not commissioned: externally peer-reviewed.

## Open access

Yes.

