## Supplementary for "The relationships between experimental task and questionnaire measures of reward/punishment sensitivity in attention deficit hyperactivity disorder (ADHD): protocol for a scoping review"

**Supplementary appendix 1: PRISMA-ScR**

**The relationships between experimental tasks and questionnaire of reward/punishment sensitivity in attention deficit hyperactivity disorder: protocol for a scoping review**

Studies from databases/registers **(n = 5,822)**

- PubMed (2,109)
- PsycINFO (2,109)
- Web of Science (1,177)

References from other sources **(n = )**

Citation searching (n = )

**Identification**

References removed **(n = 1093)**

Duplicates identified manually (n = )

Duplicates identified by Covidence (n = 1093)

Marked as ineligible by automation tools (n = )

Other reasons (n = )

Studies excluded **(n = 0)**

Studies not retrieved **(n = 0)**

Studies assessed for eligibility **(n = 0)**

Studies sought for retrieval **(n = 0)**

Studies screened **(n = 0)**

Studies excluded **(n = 0)**

**Screening**

**Included**

Studies included in review **(n = 0)**

Included studies ongoing **(n = 0)**

Studies awaiting classification **(n = 0)**

**Supplementary appendix 2-1: Search strategy in PubMed**

This scoping review’s search strategy was based on the following table in PubMed. The first part of each block included Medical Subject Headings (MeSH) terms (1 & 4 & 7). Using All fields, the terms related to MeSH or suited for the scoping review purpose were used to cover the wide range of preliminary research in other parts of each block (2 & 5 & 8 & 9 & 10).

| **Title** | The relationships between experimental tasks and questionnaire of reward/punishment sensitivity in attention deficit hyperactivity disorder: protocol for a scoping review |
| --- | --- |
| **Question** | 1. Examine which aspects of hypothesized altered reward and punishment sensitivity correspond to constructs measured by existing questionnaires. 2. Characterize the relationships between ADHD symptomatology and reward and punishment sensitivity as measured by existing questionnaires. 3. Evaluate the consistency between the altered reward and punishment sensitivity as measured by existing questionnaires and in experimental tasks. |
| **Database** | PubMed |
| **Date** | 2023/07/17 |
| **Searcher** | NN/MO |

| **#** | **Search terms** | **Items found** |
| --- | --- | --- |
| ***Population: People who have diagnosis or tendency of ADHD symptoms*** | | |
| 1 | "attention deficit disorder with hyperactivity"[MeSH Terms] | 34,672 |
| 2 | "attention deficit disorder with hyperactivity"[All Fields] OR "adhd"[All Fields] OR "add"[All Fields] OR "attention deficit hyperactivity disorder*"[All Fields] OR "hyperkinetic syndrome"[All Fields] OR "addh"[All Fields] OR "attention deficit disorder*"[All Fields] OR "minimal brain dysfunction"[All Fields] | 134,893 |
| ***Combined sets*** | | |
| 3 | **1 OR 2** | 134,893 |
| ***Measurements: Sets of questions to obtain information from participants about a topic of interest.*** | | |
| 4 | "surveys and questionnaires"[MeSH Terms] OR "self report"[MeSH Terms] OR "psychometrics"[MeSH Terms] | 1,258,002 |
| 5 | "measurement*"[All Fields] OR "scale*"[All Fields] OR "questionnaire*"[All Fields] OR "survey*"[All Fields] OR "experiment*"[All Fields] OR "task*"[All Fields] | 6,887,690 |
| ***Combined sets*** | | |
| 6 | **4 OR 5** | 7,294,063 |
| ***The concepts of measurement: Assessing the reward and punishment sensitivity.*** | | |
| 7 | "reinforcement, psychology"[MeSH Terms] OR "punishment"[MeSH Terms] OR "motivation"[MeSH Terms] OR "decision making"[MeSH Terms] OR "pleasure"[MeSH Terms] OR “reinforcement schedule”[MeSH Terms] | 454,374 |
| 8 | "reward sensitivity"[All Fields] OR "punishment sensitivity"[All Fields] OR "sensitivity to reward"[All Fields] OR "sensitivity to punishment"[All Fields] OR "sensitivity to reward and punishment"[All Fields] OR "partial reinforcement"[All Fields] OR "continuous reinforcement"[All Fields] OR “reward responsiveness”[All Fields] OR “reward seeking” [All Fields] | 3,591 |
| 9 | "behavioral inhibition system"[All Fields] OR "behavioral activation system"[All Fields] OR "behavioral approach system"[All Fields] OR "reinforcement sensitivity theory"[All Fields] | 594 |
| 10 | "generalized reward"[All Fields] OR "sensitivity to punishment and sensitivity to reward"[All Fields] OR "fawcett clark pleasure"[All Fields] OR "snaith hamilton pleasure"[All Fields] OR "chapman anhedonia"[All Fields] OR "specific loss of interest and pleasure"[All Fields] OR "temporal experience of pleasure"[All Fields] OR "anticipatory and consummatory interpersonal pleasure"[All Fields] OR "rewarding events"[All Fields] OR "motivation and pleasure"[All Fields] OR "dimensional anhedonia"[All Fields] OR “quick delay”[All Fields] | 559 |
| ***Combined sets*** | | |
| 11 | **7 OR 8 OR 9 OR 10** | 456,031 |
| 12 | **3 AND 6 AND 11** | 2,600 |
| 13 | **12 NOT (“systematic review”[Title] OR “meta-analysis”[Title]) AND ((excludepreprints[Filter]) AND (humans[Filter]) AND (english[Filter]))** | 2,109 |

**Supplementary appendix 2-2: Search strategy in Web of Science**

Based on the search strategy of PubMed, the search terms were decided in Web of Science.

| **Title** | The relationships between experimental tasks and questionnaire of reward/punishment sensitivity in attention deficit hyperactivity disorder: protocol for a scoping review |
| --- | --- |
| **Question** | 1. Examine which aspects of hypothesized altered reward and punishment sensitivity correspond to constructs measured by existing questionnaires. 2. Characterize the relationships between ADHD symptomatology and reward and punishment sensitivity as measured by existing questionnaires. 3. Evaluate the consistency between the altered reward and punishment sensitivity as measured by existing questionnaires and in experimental tasks. |
| **Database** | Web of Science |
| **Date** | 2023/07/17 |
| **Searcher** | NN/MO |

| **#** | **Search terms** | **Items found** |
| --- | --- | --- |
| ***Population: people who have diagnosis or tendency of ADHD symptoms*** | | |
| 1 | (((((((((ALL=("attention deficit disorder with hyperactivity")) OR ALL=("adhd")) OR ALL=("add")) OR ALL=("attention deficit hyperactivity disorder")) OR ALL=("hyperkinetic syndrome")) OR ALL=("adhd")) OR ALL=("attention deficit disorder")) OR ALL=("attention deficit hyperactivity disorders")) OR ALL=("attention deficit disorders")) OR ALL=("minimal brain dysfunction") | 198,301 |
| ***Measurements: Sets of questions to obtain information from participants about a topic of interest.*** | | |
| 2 | (((((((((((((ALL=("questionnaire")) OR ALL=("questionnaires")) OR ALL=(measurement)) OR ALL=("measurements")) OR ALL=("scale")) OR ALL=("scales")) OR ALL=("self report")) OR ALL=("self-report")) OR ALL=("psychometric")) OR ALL=("psychometrics")) OR ALL=("experiment")) OR ALL=("experiments")) OR ALL=("task")) OR ALL=("tasks") | 8,916,679 |
| ***The concepts of measurement: Assessing the reward and punishment sensitivity.*** | | |
| 3 | (((((ALL=("reinforcement" )) OR ALL=("punishment")) OR ALL=("motivation")) OR ALL=("decision make")) OR ALL=("decision making")) OR ALL=("pleasure") | 761,668 |
| 4 | (((((((((ALL=("reward sensitivity")) OR ALL=("punishment sensitivity")) OR ALL=("sensitivity to reward")) OR ALL=("sensitivity to punishment")) OR ALL=("sensitivity to reward and punishment")) OR ALL=("partial reinforcement")) OR ALL=("continuous reinforcement")) OR ALL=("reinforcement schedule")) OR ALL=(“reward responsiveness”)) OR ALL=(“reward seeking”) | 5,464 |
| 5 | (((ALL=("behavioral inhibition system")) OR ALL=("behavioral activation system")) OR ALL=("behavioral approach system")) OR ALL=(“reinforcement sensitivity theory”) | 1,269 |
| 6 | (((((((((((ALL=("generalized reward")) OR ALL=("sensitivity to punishment and sensitivity to reward")) OR ALL=("fawcett clark pleasure")) OR ALL=("snaith hamilton pleasure")) OR ALL=("chapman anhedonia")) OR ALL=("specific loss of interest and pleasure")) OR ALL=("temporal experience of pleasure")) OR ALL=("anticipatory and consummatory interpersonal pleasure")) OR ALL=("rewarding events")) OR ALL=("motivation and pleasure")) OR ALL=("dimensional anhedonia")) OR ALL=(“quick delay”) | 599 |
| ***Combined sets*** | | |
| 7 | **3 OR 4 OR 5 OR 6** | 763,684 |
| 8 | **1 AND 2 AND 7** | 2,766 |
| 9 | **8 NOT (TI=("systematic review") OR TI=("meta-analysis")) and English (Languages) and Article or Early Access (Document Types)** | 2,536 |

**Supplementary appendix 2-3: Search strategy**

| **Title** | The relationships between experimental tasks and questionnaire of reward/punishment sensitivity in attention deficit hyperactivity disorder: protocol for a scoping review |
| --- | --- |
| **Question** | 4. Examine which aspects of hypothesized altered reward and punishment sensitivity correspond to constructs measured by existing questionnaires. 5. Characterize the relationships between ADHD symptomatology and reward and punishment sensitivity as measured by existing questionnaires. 6. Evaluate the consistency between the altered reward and punishment sensitivity as measured by existing questionnaires and in experimental tasks. |
| **Database** | PsycINFO |
| **Date** | 2023/07/17 |
| **Searcher** | NN |

| **#** | **Search terms** | **Items found** | |
| --- | --- | --- | --- |
| ***Population: People who have diagnosis or tendency of ADHD symptoms*** | | | |
| 1 | attention deficit disorder with hyperactivity/ or attention deficit disorder/ | 31,790 | |
| 2 | "attention deficit disorder with hyperactivity" or "attention deficit disorder*" or "attention deficit hyperactivity disorder*" or "add" or "adhd" or "addh" or "hyperkinetic syndrome" or "minimal brain dysfunction" | 77,644 | |
| ***Combined sets*** | | | |
| 3 | **1 or 2** | | 77,644 |
| ***Measurements: Sets of questions to obtain information from participants about a topic of interest.*** | | | |
| 4 | questionnaires/ or surveys/ or measurement/ or psychometrics/ or likert scales/ or rating scales/ or self-report/ | 191,153 | |
| 5 | "measurement*" or "scale*" or "questionnaire*" or "survey*" or "experiment*" or "task*" | 2,205,941 | |
| ***Combined sets*** | | | |
| 6 | **4 or 5** | 2,220,013 | |
| ***The concepts of measurement: Assessing the reward and punishment sensitivity.*** | | | |
| 7 | reinforcement/ or punishment/ or motivation/ or decision making/ or pleasure/ | 174,956 | |
| 8 | reward sensitivity/ or reinforcement schedules/ or punishment/ | 15,331 | |
| 9 | "punishment sensitivity" or "sensitivity to punishment" or "sensitivity to reward and punishment" or "reward responsiveness" or "reward seeking" | 2062 | |
| 10 | behavioral inhibition system/ or behavioral activation system/ | 872 | |
| 11 | "reinforcement sensitivity theory" | 422 | |
| 12 | "generalized reward" or "sensitivity to punishment and sensitivity to reward" or "fawcett clark pleasure" or "snaith hamilton pleasure" or "chapman anhedonia" or "specific loss of interest and pleasure" or "temporal experience of pleasure" or "anticipatory and consummatory interpersonal pleasure" or "rewarding events" or "motivation and pleasure" or "dimensional anhedonia" or "quick delay" | 1540 | |
| ***Combined sets*** | | | |
| 13 | **7 or 8 or 9 or 10 or 11 or 12** | 183,599 | |
| ***Combined sets*** | | | |
| 14 | **3 and 6 and 13** | 1290 | |
| 15 | **(limit 14 to (English and human)) not ("systematic review" or "meta-analysis").ti.** | 1177 | |

**Supplementary appendix 3: Draft Data Collection Items**

- Source

・Year

・Author

・Title

・Citation Number

- Eligibility

・Reason for exclusion

・Reason for inclusion

- Methods

・Design: intervention, experimental task, questionnaire

・Purpose/Study Question

- Population Characteristics

・Age

・Number

・Number of Group

・Sex

・Diagnosis/Symptoms

・Country

・Co-morbidity

・Socio-demographics/Ethnicity/Other

- Measurements for the reinforcement and punishment sensitivity (Outcomes)

・Name of Measurement (experimental tasks & scales)

・Developer

・Development Year

・Definition of Measurement (experimental tasks & scales)

- Results

・Outcomes

・Effect size, CI, *p*-values

- Others

・Funding

・Key conclusions

・Comments

・Relevant references
